# Physiology-Based Electrocardiographic Criteria for Left Bundle Branch Capture

**DOI:** 10.1101/2020.12.24.20248827

**Authors:** Marek Jastrzębski, Grzegorz Kiełbasa, Karol Curila, Paweł Moskal, Agnieszka Bednarek, Marek Rajzer, Pugazhendhi Vijayaraman

## Abstract

**Background:** During left bundle branch (LBB) area pacing, it is important to confirm that the capture of the LBB is achieved, not just the capture of only the adjacent left ventricular myocardium (LV septal capture). Our aim was to establish ECG criteria for LBB capture by analyzing ECGs with confirmed LBB capture and non-capture. We hypothesized that since LBB pacing results in physiologic depolarization of the left ventricle then the native QRS can serve as a reference for the diagnosis of LBB capture in the same patient.

**Methods:** Only patients with direct evidence of LBB capture (output-dependent or refractoriness-dependent QRS morphology transition) were included. Several QRS characteristics were compared between the native rhythm and different types of LBB area capture. Receiver-operator characteristics analysis was performed to determine the optimal V6 R-wave peak time (RWPT) cut-off for LBB diagnosis.

**Results:** A total of 357 ECG tracing (124 patients) were analyzed: 118 with native rhythm, 124 with non-selective LBB capture, 69 with selective LBB capture and 46 with LV septal capture. Our hypotheses that during LBB capture the paced V6 RWPT (measured from QRS onset) equals the native V6 RWPT and that the paced V6 RWPT (measured from the stimulus) equals the LBB potential to V6 R-wave peak interval were positively validated. Criteria based on these rules had sensitivity and specificity of 98.0–88.2% and 85.7–95.4%, respectively. The optimal and 100% specific V6 RWPT values for differentiation between LBB capture and LV septal capture in patients with narrow QRS / right bundle branch block were 83 ms and 74 ms, respectively; while in patients with left bundle branch block/asystole/ventricular escape the optimal and 100% specific V6 RWPT values were 101 ms and 80 ms, respectively.

**Conclusions:** Novel criteria for LBB capture were developed and optimal V6 RWPT cut-offs were determined.

**What this study adds:**

- We showed that LBB pacing truly reproduce the physiological depolarization of the left ventricle since the paced V6 RWPT equals the native conduction V6 RWPT.
- Individualized LBB capture criteria, that use the native QRS as a reference, were developed.
- The optimal V6 RWPT values for differentiation between LBB capture and LV septal capture were determined, separately for patients with healthy and diseased LBB.

## Introduction

Left bundle branch (LBB) pacing is rapidly developing into a promising pacing technique with potential for application in both conventional pacing and heart failure patients.^1,3^ During LBB pacing, it is important to confirm that the capture of the LBB is achieved, not just the capture of only the adjacent left ventricular septal myocardium (LV septal pacing).

Although a few ECG criteria (QRS duration, V6 R-wave peak time (RWPT)) for LBB capture were proposed to differentiate between non-selective (ns) LBB pacing and LV septal pacing, these are arbitrary and were never validated, with unknown sensitivity and specificity.^1-5^

Using dynamic ECG maneuvers with output-dependent and refractoriness-dependent QRS morphological changes as the ‘gold standard’ for LBB capture diagnosis (Supplemental Figures 1 and 2),^1,4^ it should be possible to precisely characterize various types of LBB area paced QRS complexes. This could serve to develop/validate ECG criteria for differentiation between LBB capture vs. LV septal capture.

Moreover, we believe that since conduction system pacing results in physiological depolarization of the left ventricle, then the native QRS complex parameters in a patient could be used as a reference to assess if the paced QRS complex in the same patient is compatible with physiological depolarization of the left ventricle. On the basis of this, the following specific research hypotheses were formulated:

1. Capture of the LBB can be diagnosed when the paced V6 RWPT (measured from the QRS onset) equals the V6 RWPT during native non-LBBB rhythm - Figure 1.
2. Capture of the LBB can be diagnosed when the paced V6 RWPT (measured from the stimulus) equals the LBB potential to the V6 R-wave peak interval during native non-LBBB rhythm - Figure 2.
3. Capture of the LBB in a patient with LBBB can be diagnosed when the paced V6 RWPT is shorter than the native V6 intrinsicoid deflection time (IDT) by more than the transseptal conduction time (TCT) - Figure 3.

**Figure 1.**
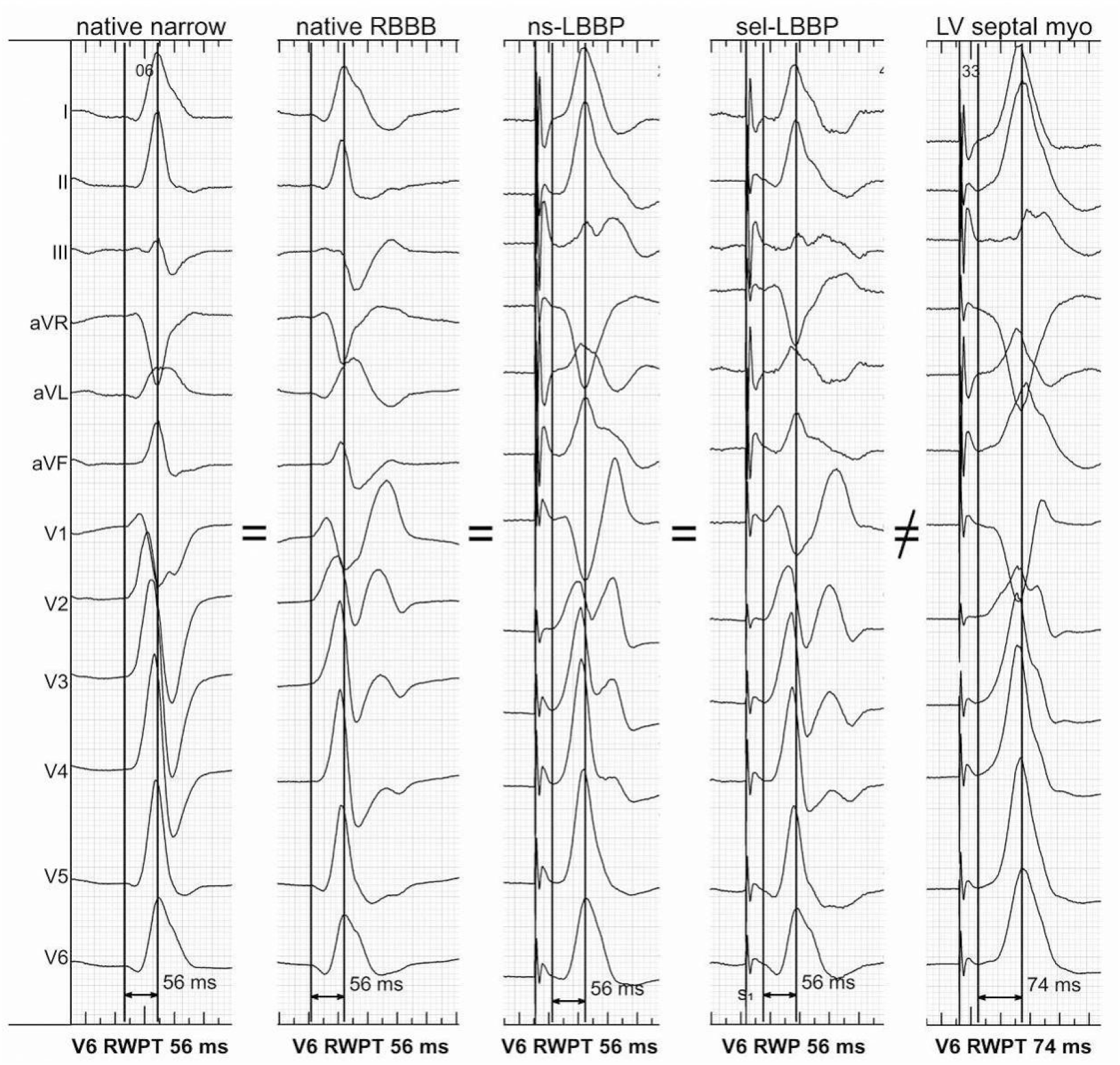
Five types of QRS complexes were obtained in the same patient. The R wave peak time in lead V6 (V6 RWPT) remained the same during the activation of the left bundle branch (LBB) via either intrinsic rhythm with narrow QRS or right bundle branch block (RBBB), or via pacing, either with non-selective LBB pacing (ns-LBBP) or selective LBB pacing (sel-LBBP). However, the V6 RWPT was evidently longer without the LBB capture, i.e. during left ventricular septal myocardial (LV septal myo) pacing.

**Figure 2.**
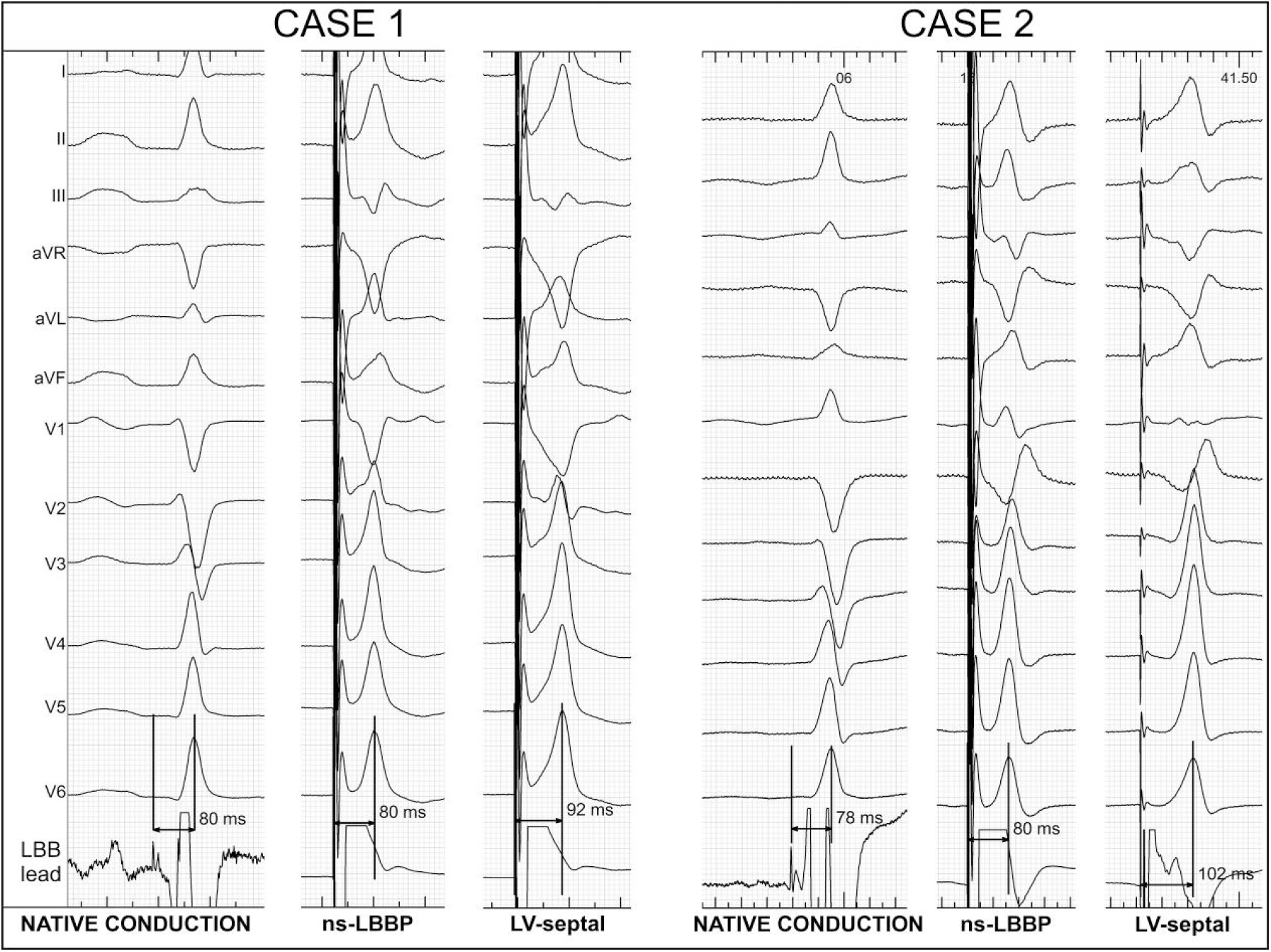
During non-selective capture of the left bundle branch (ns-LBBP), the interval from the stimulus to the R wave peak equaled the interval from the left bundle branch potential to the R wave peak during native conduction. During loss of left bundle branch capture, resulting in only left ventricular septal myocardial pacing (LV-septal), the interval from the stimulus to the R wave peak was longer.

**Figure 3.**
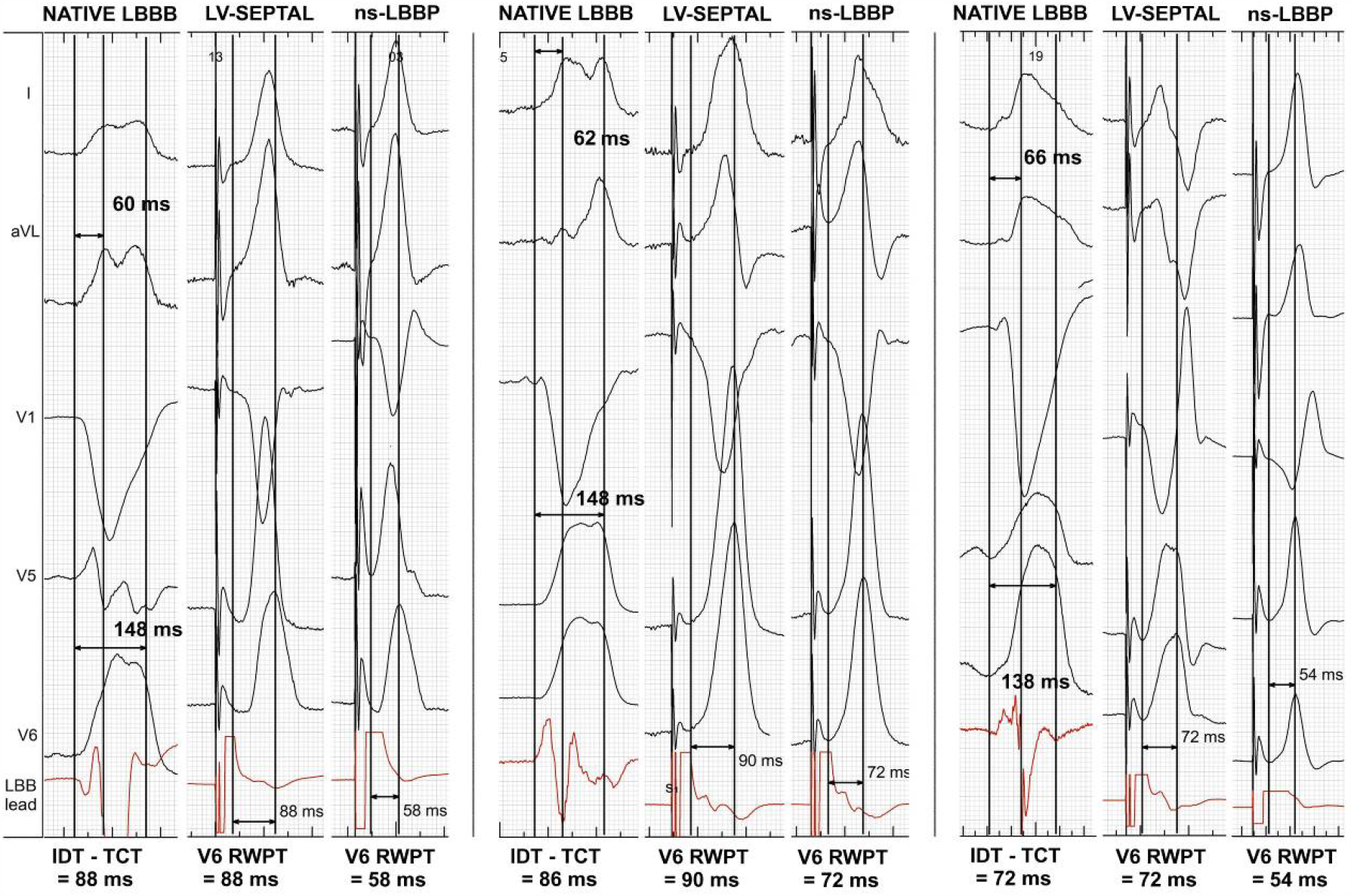
Three cases of the left bundle branch block (LBBB) corrected with non-selective left bundle branch pacing (ns-LBBP). The V6 RWPT during ns-LBBP was shorter than the V6 RWPT during left ventricular septal myocardial pacing (LV septal) that served as a reference for diagnosis of LBB capture. The V6 RWPT of LV septal pacing can be approximated by subtracting the transseptal conduction time (TCT) from the intrinsicoid deflection time (IDT).

## Aim

Our aim was to establish new ECG criteria for diagnosis of LBB capture and to determine diagnostically optimal cut-off for V6 RWPT.

## Methods

The data that support the findings of this study are available from the corresponding author upon reasonable request.

### Population

Patients who underwent LBB pacing device implantation for bradycardia and/or heart failure indications in two centers were screened. The LBB implantation procedure was described by us and others elsewhere.^1-7^ In order to develop LBB capture criteria, only patients with direct evidence of capture of the LBB were included in the study. Such a situation was considered to be the case when there was QR/rSR’ paced QRS morphology in lead V1, with at least one of the following three conditions fulfilled:

1. Transition from ns-LBB to selective (s)-LBB capture was observed during decrease in pacing output.
2. Transition from ns-LBB to LV septal pacing was observed during decrease in pacing output.
3. Transition from ns-LBB to s-LBB capture was observed during programmed stimulation and/or rapid pacing.

### Definitions and measurements

In every studied patient, each available paced QRS type (s-LBB, ns-LBB and LV septal) as well as QRS during the native rhythm – supraventricular and/or escape – was measured. The following QRS characteristics were obtained:

1. Global QRS duration
2. V6 RWPT
3. LBB potential to QRS onset interval
4. LBB potential to V6 R-wave peak interval
5. V6 IDT – only in patients with LBBB
6. TCT – only in patients with LBBB

Global QRS duration, as well as V6 RWPT, was measured using two methods: from the pacing artifact and from the QRS onset, i.e. without the pacing artifact and the initial latency (Figure 1). To allow for some imprecisions in measurement, a 10-ms difference between the native conduction interval and the corresponding interval during pacing was permitted.

In patients with LBBB, the V6 IDT was measured instead of the V6 RWPT, because in LBBB morphology the peak time and the IDT are usually not equivalent. The IDT was measured from the earliest QRS onset in any surface lead (global method), not to the point of the highest amplitude, but to the end of the slur/plateau in QRS, that is to the beginning of the final rapid downsloping phase of R wave in lead V6 (Figure 3). In cases with monophasic QRS in V6 (a smooth descent from the peak to the end), the beginning of the final rapid downsloping phase in V5, I, and/or aVL were used.

TCT was measured using the endocardial signal from the pacing lead implanted in the LBB area and the surface ECG. TCT was considered equal to the time interval from the earliest QRS onset in any surface lead to the endocardial indication of the arrival of the depolarization wavefront to the LBB area. This was indicated by the first rapid change of endocardial signal polarity or velocity – usually it is the first spiky peak/nadir, but occasionally the first rapid acceleration of the endocardial signal (Figure 3). To avoid mistakes in choosing the right component of the often multicomponent endocardial signal, it has to be coincident (+/- 15 ms) with the very beginning of the first notch/slur/plateau in the lateral leads, preferably leads I and aVL, as their vectors record the septal activation more globally than the unipolar V6 that is far from the septum (Figure 3).

Every implantation procedure was recorded on the digital electrophysiological system (LabsystemPRO, Boston Scientific, USA). To ensure high precision, the measurements were performed using all 12 surface ECG leads recorded simultaneously, digital calipers, fast sweep speed (200 mm/s), and appropriate signal augmentation. At least three QRS complexes were measured and their values averaged.

All patients gave written informed consent for participation in this study and the Institutional Bioethical Committee approved the study protocol.

### Statistical analysis

Continuous variables are presented as means and standard deviations. The distribution of the QRS and lead V6 RWPT was estimated by the kernel method. Categorical variables are presented as percentages. Differences between groups were assessed using the Fisher exact test for a 2 × 2 table or Student’s t-test, as appropriate. The performance of binary decision rules was described using sensitivity (SN) and specificity (SP). The performance of the QRS duration and V6 RWPT in discriminating between ns-HB and RV pacing was assessed using the receiver operating characteristic curve (ROC). Statistical analyses were performed in R. P-values < 0.05 were considered statistically significant.

## Results

### Population

A total of 468 patients with LBB area pacing from two centers were screened, and 124 cases with certain diagnosis of LBB capture were included in the final analysis. In the excluded patients, the LBB diagnosis was based on some arbitrary criteria,^1,5^ but could not be confirmed with dynamic QRS morphological changes. The basic clinical characteristics of the included patients is presented in Table 1. A total of 357 ECG tracings were analyzed: 118 with native rhythm, 124 with ns-LBB capture, 69 with s-LBB capture and 46 with LV septal capture.

**Table 1.** Basic characteristics of the studied group (n = 124)

|  |  |
| --- | --- |
| Age [years] | 74.7 ± 11.4 |
| Male gender | 71 (57.3%) |
| Pacing indication [n]: |  |
| • Sick sinus syndrome | 32 (25.8%) |
| • Atrioventricular block | 41 (33.1%) |
| • Atrial fibrillation with bradycardia | 13 (10.5%) |
| • Heart failure | 38 (30.6%) |
| Comorbidities [n]: |  |
| • Diabetes mellitus | 49 (39.5%) |
| • Coronary heart disease | 51 (41.1%) |
| • Heart failure | 59 (47.6%) |
| • Hypertension | 99 (79.8%) |
| • Severe valvular disease | 16 (12.9%) |
| Left ventricular ejection fraction [%] | 46.2 ± 15.5 |
| Left ventricular end-diastolic dimension [mm] | 53.0 ± 8.7 |
| Native QRS duration [ms] | 133.6 ± 34.8 |
| Native QRS type: |  |
| • Narrow | 39 (31.6%) |
| • LAFB | 5 (4.0%) |
| • RBBB | 13 (10.5%) |
| • RBBB + LAFB/LPFB | 15 (12.1%) |
| • NIVCD | 15 (12.1%) |
| • LBBB | 18 (14.5%) |
| • Escape with RBBB-type QRS | 13 (10.5%) |
| • Escape with LBBB-type QRS | 2 (1.6%) |
| • Complete asystole (paced) | 4 (3.2%) |

3 RBBB – right bundle branch block; LAH – left anterior fascicular block; LPH – left posterior 4 fascicular block; LBBB – left bundle branch block; NIVCD – non-specific intraventricular 5 conduction disturbance.

### Novel criteria for diagnosis of LBB capture in non-LBBB patients

In the subgroup of 100 non-LBBB patients (after excluding LBBB, LBBB-type escape and asystole cases), a total of 290 ECG tracings were analyzed: 100 with native rhythm, 100 with ns-LBB capture, 55 with s-LBB capture and 35 with LV septal capture.

The correlation between the native V6 RWPT and the ns-LBB paced V6 RWPT (without latency) was strong: r =0.73, p <0.01 (Figure 4). These two measures were nearly equal, with an average difference of 2.7 ms (47.7 ±9.6 ms vs. 45.0 ±10.4 ms). The average latency between the stimulus and the QRS onset was 31.3 ± 4.6 ms.

**Figure 4.**
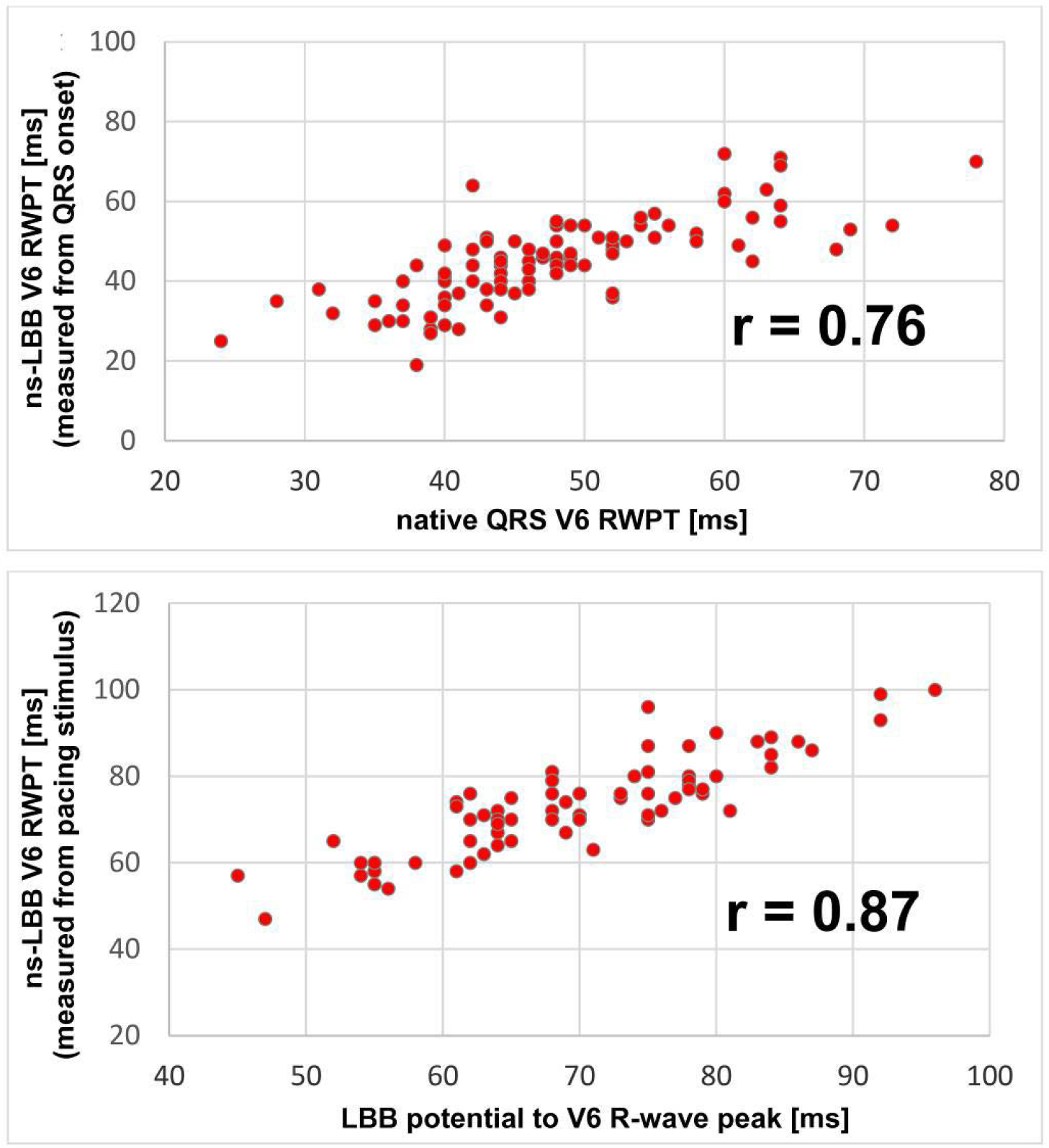
Correlation between native QRS parameters (x) and paced QRS parameters (y). <u>Upper panel</u>: correlation between the native and the non-selective (ns) left bundle branch (LBB) paced V6 R wave peak time (RWPT) measured from the QRS onset. <u>Lower panel:</u> correlation between LBB potential to R-wave peak in V6 and V6 RWPT measured from the pacing stimulus.

Similarly, the correlation between the LBB potential to V6 R-wave peak interval and the V6 RWPT measured for the stimulus was strong: r =0.87, p <0.01 (Figure 4). The average difference between them was 3.5 ms (69.9 ±10.7 ms vs. 73.4 ±11.0 ms). The average potential to QRS interval was 22 ±7.0 ms.

The criterion of “paced V6 RWPT (measured from QRS onset) ≤ native V6 RWPT (+10ms)” correctly diagnosed ns-LBB capture in 98/100 patients with ns-LBB capture and misdiagnosed ns-LBB in 3/35 patients with LV septal pacing. This resulted in SN of 98.0% and SP of 85.7% for the diagnosis of LBB capture.

In this subgroup, 68 (68%) patients had LBB potential recorded. The criterion of “paced V6 RWPT (measured from stimulus) ≤ LBB potential to V6 R-wave peak interval (+10 ms)” correctly diagnosed ns-LBB capture in 60/68 of patients with ns-LBB capture and misdiagnosed ns-LBB capture in 1/22 patients with LV septal pacing. This resulted in SN of 88.2% and SP of 95.4% for the diagnosis of LBB capture.

There was no difference between ns-LBB and s-LBB paced QRS with regard to V6 RWPT when measured from the stimulus or from the QRS onset: 74.7 ±12.0 ms vs. 74.4 ±13.0 ms (p = 0.63) and 43.6 ±9.5 ms vs. 43.2 ±10.8 ms (p = 0.67), respectively.

### Novel criteria for diagnosis of LBB capture in LBBB patients

A total of 18 patients with LBBB were analyzed. In 12/18 patients transition to s-LBB capture was observed, in 8/18 transition to LV septal capture was observed, and in 12/18 a selective response during programmed stimulation was observed (in several patients two or all three phenomena were present). In patients with LBBB, the average TCT was 63.0 ± 8.2 ms and the average IDT was 128 ± 16.8 ms. The LV septal paced V6 RWPT (measured from QRS onset) was closely approximated by the difference IDT – TCT (71.4 ±11.4 ms vs. 66.0 ±12.4, ms, p = 0.19). The ns-LBB paced V6 RWPT (measured from QRS onset) was, in 14/18 cases, shorter by > 10 ms than the difference IDT – TCT, while the LV septal paced V6 RWPT (measured from QRS onset) in 8/18 was equal to, or longer or shorter by less than 10 ms than, the difference IDT – TCT. Consequently, the criterion of “paced V6 RWPT +10 ms < (IDT – TCT)” had SN and SP, for the diagnosis of LBB capture in patients with LBBB, of 77.8% and 100%, respectively.

In 4/18 LBBB patients, intraprocedural mechanical trauma to the right bundle branch resulted in a complete transient atrioventricular block and emergence of an escape rhythm of RBBB morphology. In all these cases, the criterion of “paced V6 RWPT (measured from QRS onset) ≤ native V6 RWPT (+10ms)” was fulfilled.

### Distribution of V6 RWPT values during different types of capture and optimal cut-offs

There was a difference in observed V6 RWPT values between patients with normal LBB conduction (narrow/RBBB QRS) and patients with diseased LBB (LBBB/NIVCD/escape/asystole), both when V6 RWPT was measured for the QRS onset or from the pacing stimulus: 42.5 ± 8.9 ms vs 51.5 ± 11.3 ms (p < 0.0001) and 73.3 ± 9.6 ms vs 84.2 ± 13.5 ms (p < 0.0001), respectively.

The distribution of V6 RWPT (measured from the pacing stimulus) in narrow QRS/RBBB patients, during ns-LBB and LV septal capture, together with the ROC curves, is presented in Figure 5. The average increase in V6 RWPT with loss of LBB capture was 20.0 ± 6.4 ms. The optimal V6 RWPT value for differentiation between LBB capture and LV septal capture was 83 ms (SN of 84.7% and SP of 96.3%). V6 RWPT value of <74 ms had SP for LBB capture diagnosis of 100%.

**Figure 5.**
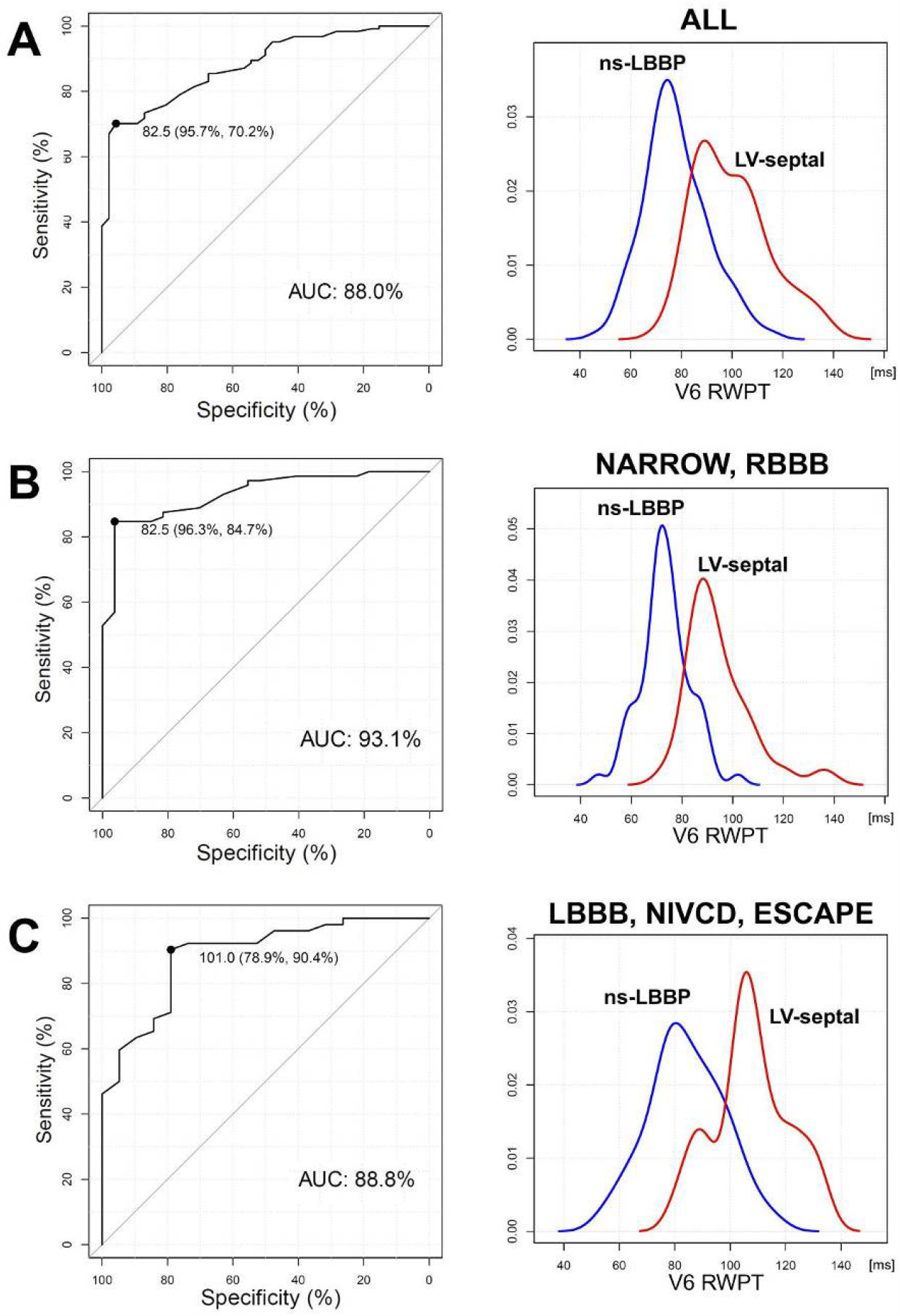
Receiver operating characteristic curves for lead V6 RWPT and the distribution of V6 RWPT values during non-selective left bundle branch pacing (ns-LBBP) and left ventricular septal myocardial pacing (LV septal). <u>Panel A</u>: data for the whole studied group. <u>Panel B:</u> data for the patients with narrow QRS or right bundle branch block (RBBB) QRS. <u>Panel C</u>: data for patients with left bundle branch block (LBBB), non-specific intraventricular conduction disturbance (NIVCD) or escape rhythm/asystole (ESCAPE). In all cases, the V6 RWPT was measured from the stimulus onset.

The distribution of V6 RWPT (measured from the pacing stimulus) in patients with LBBB/NIVCD/escape/asystole, during ns-LBB and LV septal capture, together with the ROC curves, is presented in Figure 5. The average increase in V6 RWPT with loss of LBB capture was 21.2 ± 7.5 ms. The optimal V6 RWPT value for differentiation between LBB capture and LV septal capture was 101 ms (SN of 90.4% and SP of 78.9%). V6 RWPT value of ≤ 80 ms had specificity for LBB capture diagnosis of 100%.

### QRS duration

The average QRS durations for native QRS, ns-LBB QRS, s-LBB QRS and LV septal QRS are presented in Tables 1 and 2. There was a nearly complete overlap of QRS duration values between different ns-LBB capture and LV septal pacing, eliminating QRS duration as a useful parameter for the diagnosis of LBB capture (Figure 6).

**Table 2.**
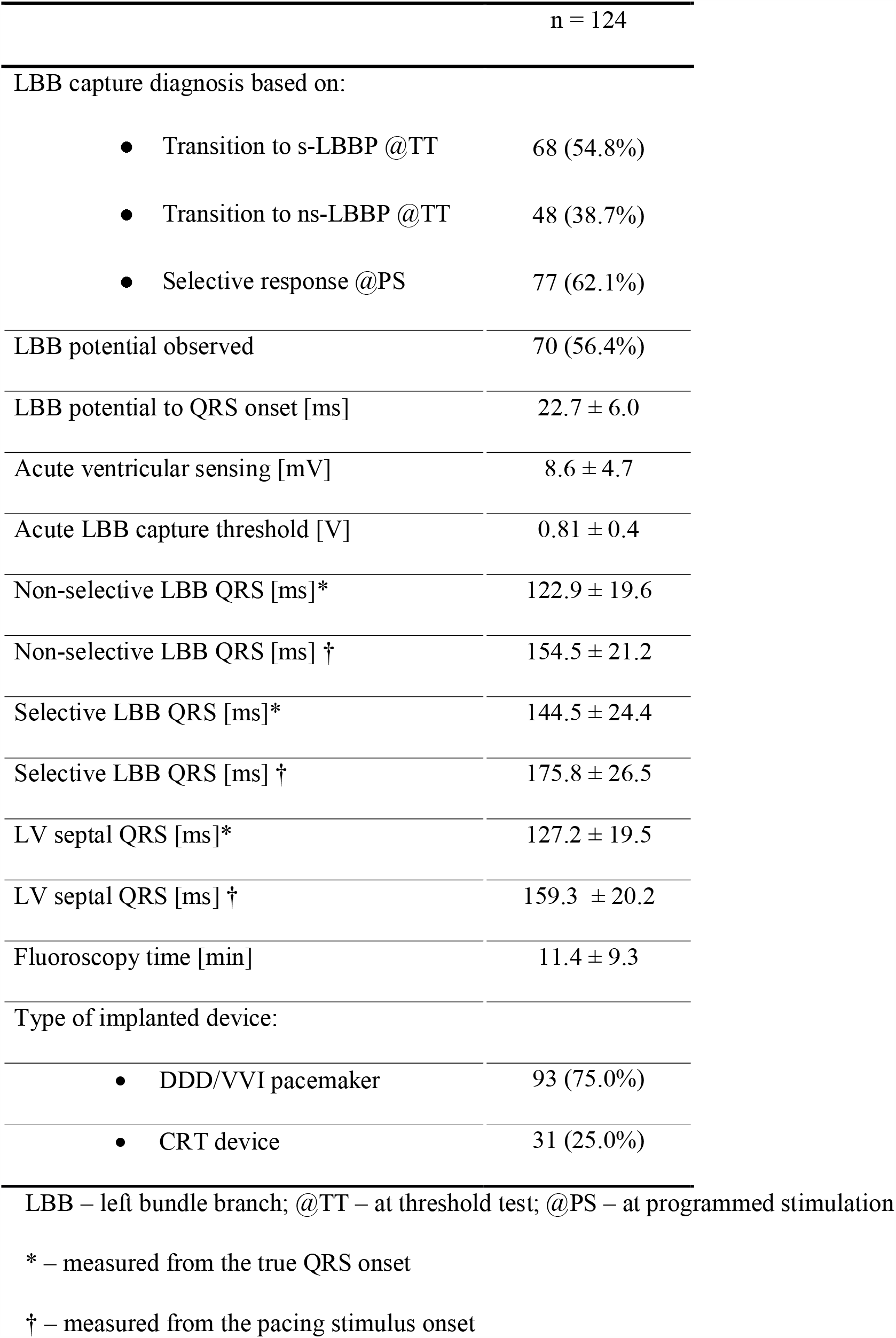
Pacing- and procedure-related characteristics

**Figure 6.**
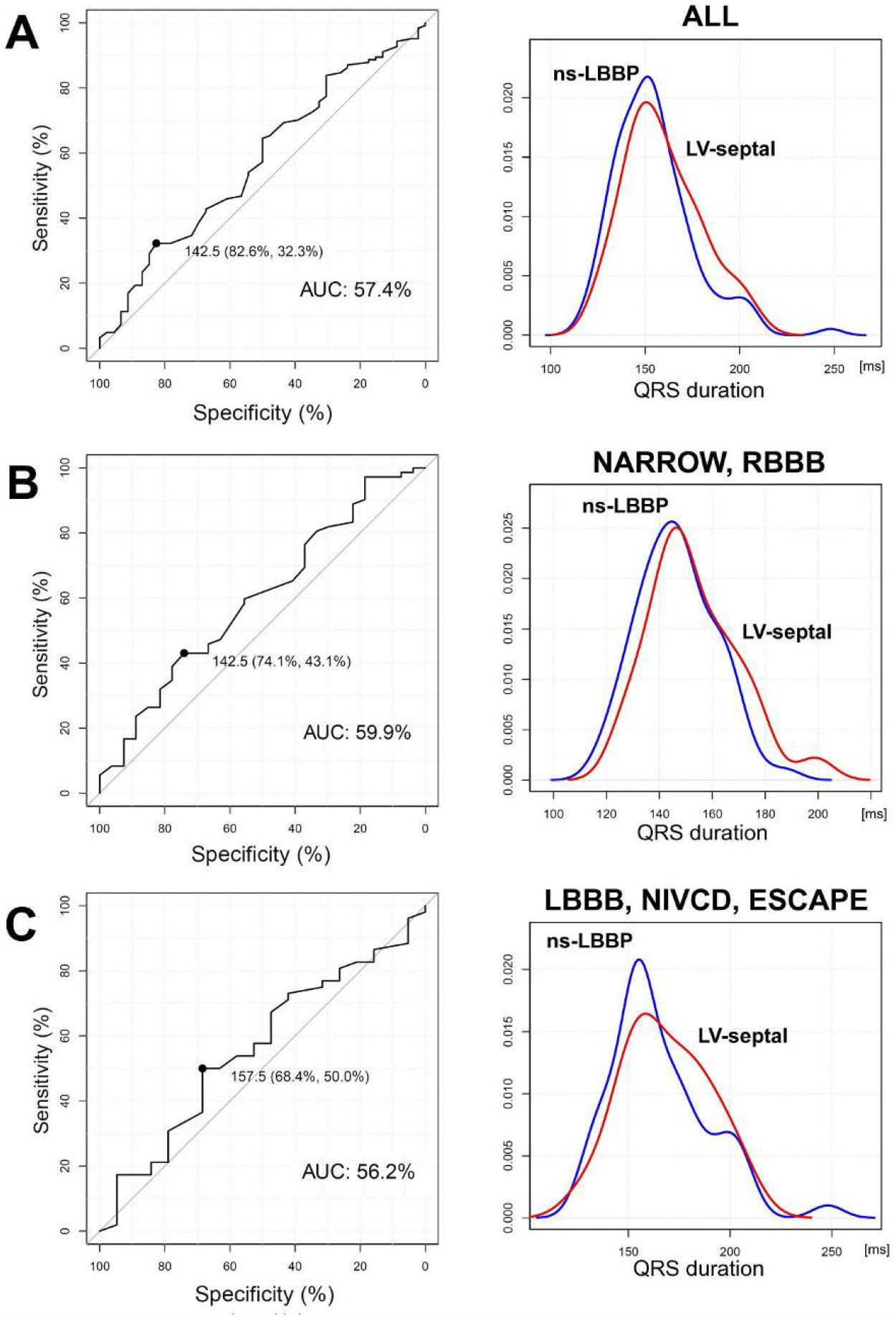
Receiver operating characteristic curves for lead QRS duration and the distribution of QRS duration values during non-selective left bundle branch pacing (ns-LBBP) and left ventricular septal myocardial pacing (LV septal). <u>Panel A</u>: data for the whole studied group. <u>Panel B:</u> data for the patients with narrow QRS or right bundle branch block (RBBB) QRS. <u>Panel C</u>: data for patients with left bundle branch block (LBBB), non-specific intraventricular conduction disturbance (NIVCD) or escape rhythm/asystole (ESCAPE). In all cases, QRS was measured from the stimulus onset.

## Discussion

The major finding of this study is that LBB capture resulted in the maintenance or restoration of the physiological activation time of the lateral wall of the LV. This was expressed by the equality of the paced V6 RWPT to the V6 RWPT during native LBB activation, and allowed us to formulate novel criteria for LBB capture.

The second important result of the study is the establishment of the optimal cut-off values of V6 RWPT for diagnosis of LBB capture in patients with healthy and with diseased LBB on the basis of a sizable sample of ECGs with confirmed LBB capture/non-capture.

### Individualized paced V6 R-wave peak time for the diagnosis of LBB capture

The V6 RWPT, in case of monophasic, single peak R wave, corresponds to the intrinsicoid deflection time in lead V6. This measure represents the time it takes for the depolarization wavefront to reach the epicardial surface of the lateral wall of the LV.^8^ During normal native conduction, the V6 RWPT depends on the conduction in the LBB and not on the RBB conduction. We hypothesized, that since during LBB capture, the activation of the lateral wall of the LV follows the physiological route, then the paced V6 RWPT should be identical as V6 RWPT during native conduction. In contrast, if LBB is not captured and only LV septal myocardial capture is present, then the activation of the lateral wall of the LV should be delayed by the time it takes for the myocardial depolarization wavefront to reach the Purkinje network–myocardium interface (usually 15–25 ms) and the paced V6 RWPT should be longer than the native V6 RWPT. The individualized V6 RWPT criterion approach is applicable to patients in whom QRS resulting from native LBB conduction can be observed (i.e. any non-LBB type QRS), either of the supraventricular or even escape origin. From our experience, this is possible in > 80% of patients referred for device implantation.

The most precise application of the above principle is comparing the stimulus to V6 R-wave peak interval with LBB potential to V6 R-wave peak interval. This study showed that during LBB capture, these two intervals were nearly identical. The pacing stimulus depolarizes the LBB at the site of the recorded potential and the activation pathways to the lateral wall of the LV are the same, with the post stimulus latency in LV depolarization equal to the latency between LBB potential and QRS onset.

This method might be inaccurate when the recorded potential is not the main LBB potential but a potential from a more distal LBB subdivision. Then, the activation pathway during pacing will not be the same but will be slightly longer by the time it takes for retrograde conduction from the point of capture to the main LBB. We observed such a situation in 9% of our patients (see supplementary materials).

When the LBB is not recorded, the V6 RWPT can be measured alternatively - directly from the QRS onset rather than from the pacing stimulus, thus omitting both the initial latency and the LBB potential to QRS interval. Our results showed that LBB capture can be diagnosed very accurately using this method.

### Fixed cut-off value for V6 R-wave peak time for the diagnosis of LBB capture

We have shown previously that during His bundle pacing, the V6 RWPT values showed typical bell-curve distribution with a significant overlap of values with those of myocardial pacing.^9^ Consequently, we assumed that a similar situation might be present during LBB pacing and that a fixed V6 RWPT was unlikely to be both specific and sensitive for the diagnosis of LBB capture. The current study confirmed this (Figures 4 and 5), showing that, during LBB capture, the V6 RWPT (measured from the pacing stimulus) could be even > 100 ms long, while, very rarely, LV septal pacing could result in V6 RWPT < 75 ms. Nevertheless, if a fixed V6 RWPT criterion is preferred, then the ROC curve analysis (Figures 4 and 5) pointed to the values 83 ms and 101 ms (measured from the pacing stimulus) as having an optimal balance of specificity and sensitivity for patients with normal and diseased LBB, respectively.

If a firm intraprocedural diagnosis of LBB capture was desired, then the cut-off values of 74 ms and 80 ms were 100% specific for LBB capture in patients with normal and diseased LBB, respectively. This corroborates and specifies the cut-off values (70, 80, and 90 ms) proposed arbitrarily by others previously.^2,4-6^ We believe that the currently established firm cut-offs might have substantial practical importance for the LBB area lead implanters.

### LBB capture in patients with LBBB

We hypothesized that LV septal pacing eliminates only the transseptal conduction delay, while LBB capture also mitigates the intra-LV delay. The V6 IDT was shown to correspond with the arrival of local myocardial depolarization to the area under the lead V6.^8^ In patients with LBBB, the V6 IDT is prolonged due to the transseptal conduction delay and the non-physiological conduction within the LV. The presence of a pacing lead in the LBB area facilitates the determination of the TCT because the arrival of the depolarization wavefront to the other side of the septum can be precisely recorded. During LV septal pacing, the V6 IDT (equivalent to the paced RWPT) is shortened only by the elimination of the transseptal conduction delay. If the paced V6 RWPT is shorter than the native IDT by more than just the ‘bypassed’ TCT, then it must be related to the complete or partial restoration of the physiological intra-LV conduction, which is possible only via recruitment of the LBB by the pacing stimulus.

Apart from the above method, the LBB capture in patients with LBBB can be occasionally diagnosed using the criterion for non-LBBB patients (native V6 RWPT = paced V6 RWPT). This is possible when a narrow or RBBB-type QRS appears either during the escape rhythm (22% of cases in the current study) or possibly during high-output His bundle pacing.

### Clinical translation

There is a consensus in the electrophysiological community involved with conduction system pacing research on the urgent need to establish consistent, precise and evidence-based, rather than arbitrary, criteria for differentiating between LBB pacing and LV septal myocardial pacing. Currently, the only available methods that can firmly ascertain the diagnosis of LBB capture are invasive (based on the mapping of His–Purkinje system activity) or require dynamic maneuvers (output-dependent and programmed LBB stimulation-induced paced QRS morphology change). The applicability of these methods to LBB capture diagnosis is limited for practical reasons and/or due to their low diagnostic sensitivity. In contrast, the currently developed criteria are based on ECG analysis, are very sensitive and are also applicable during ambulatory device follow-up.

Moreover, we demonstrated that the ECG criteria for LBB capture are deeply rooted in the normal physiology of heart depolarization. Perhaps the conduction system paced QRS should be viewed/assessed using the well-known normal values and criteria developed for the assessment of a non-paced QRS complex. For example, the average paced V6 RWPT in patients with LBBB, despite LBB capture, was longer than in cases with narrow QRS or RBBB. In addition, non-physiological paced V6 RWPT values (i.e. > 60 ms when measured from the QRS onset) were observed during LBB capture nearly only in patients with LBBB, NIVCD and asystole/escape rhythm cases, i.e. patients with very diseased left ventricular conduction systems. This suggests that despite LBB capture, some residual, but possibly clinically important, intra-LV conduction disturbance remains. In patients with indications for cardiac resynchronization therapy, such persistence of QRS features of LBBB in the paced ECG could be viewed as an indication to add a coronary sinus lead to achieve a more complete correction of the LBBB physiology. If during LV septal pacing, despite the lack of LBB capture, the paced QRS shows normal V6 RWPT (i.e. < 50 ms when measured from the QRS onset), probably resulting from a very early penetration to a healthy distal LBB, then it should be viewed as a physiological pacing as well. Notably, the 100% specific cut-off values of V6 RWPT established by this study (74–80 ms when measured from stimulus) closely correspond with the known upper limit of normal values for V6 IDT (50ms) + the post-stimulus latency interval of 30 ms.

## Limitations

This study was based on data form two centers and the results might not be universally applicable due to population and implantation technique differences.

The individualized approach to LBB criteria might be limited when the beginning of the ns-LBB paced QRS cannot be determined precisely in cases with signal oversaturation post pacing stimulus extending beyond the latency interval.

It seems that during rS morphology, the V6 RWPT is shortened by cancellation forces and probably less accurately reflects the timing of the local depolarization wavefront. Consequently, discordant morphology, i.e. when there is R/Rs in V6 during pacing and rS during native rhythm, might lead to a false negative result (see supplementary material).

## Conclusions

LBB pacing is a method of physiological pacing and can be recognized in the ECG on the basis of physiology-based criteria proposed in this study. Native QRS is a powerful reference to the diagnosis of LBB capture during LBB area pacing. It is on this basis that these novel and accurate criteria for LBB capture have been developed. It seems warranted to study further if normal values developed for the non-paced QRS complex can be used to diagnose/assess normal depolarization/electrical synchrony during LBB area pacing.

## Funding

This paper was partialy supported by the Charles University Research Program Q38, Research Centre program No. UNCE/MED/002, 260530/SVV/2020

## Disclosures

Dr. Vijayaraman has received research and fellowship support as well as speaker and consultant fees from Medtronic. He has also received consultant fees from Abbott, Biotronik, Eaglepoint LLC and Boston Scientific. Dr. Jastrzebski has received consultant fees and other honoraria from Medtronic. Dr. Curila has received consultant fees and proctoring honoraria from Medtronic. Dr. Moskal has receivedc consultant fees from Medtronic. The other authors report no conflicts.

## Notes

### Competing Interest Statement

The authors have declared no competing interest.

### Author Declarations

Jagiellonian University Bioethical committee

