## Supplemental Figures for "Physiology-Based Electrocardiographic Criteria for Left Bundle Branch Capture"

**Supplementary Results**

Post-hoc analysis of diagnostic failures

1. *Paced ns-LBB V6 RWPT > native V6 RWPT*

Paced V6 RWPT was longer (> 10 ms) than native V6 RWPT in 2/100 (0.9%) patients with non-LBBB native QRS. In both these cases, a discordant V6 morphology was present, that is, there was rS morphology during native rhythm vs. R morphology during pacing. In the correctly diagnosed patients, such a situation was much less common (8/98; 8.2%; p = 0.009).

2. *Paced ns-LBB V6 RWPT (measured from the stimulus) > LBB potential to V6 R-wave peak interval*

In 9 (13.0%) patients, the paced ns-LBB V6 RWPT (measured from the pacing stimulus) was longer (> 10 ms) than the LBB potential to V6 R-wave peak interval. The LBB potential to QRS onset interval was shorter in this subgroup than in the correctly diagnosed patients (18.7 ±3.7 ms vs. 23.5 ±5.3ms, p = 0.017, respectively); in 6/9 patients, this interval was < 20 ms.

Analysis of patients with non-physiological V6 RWPT during LBB capture

During LBB capture, V6 RWPT value (measured from real QRS onset) > 60 ms was seen in 12/124 patients. All 12 cases had diseased His-Purkinje system: 3 with LBBB, 4 with NIVCD, 4 with ventricular escape/asystole, and one with RBBB, left anterior fascicular block and QRS of 188 ms.

**Supplementary Figures**

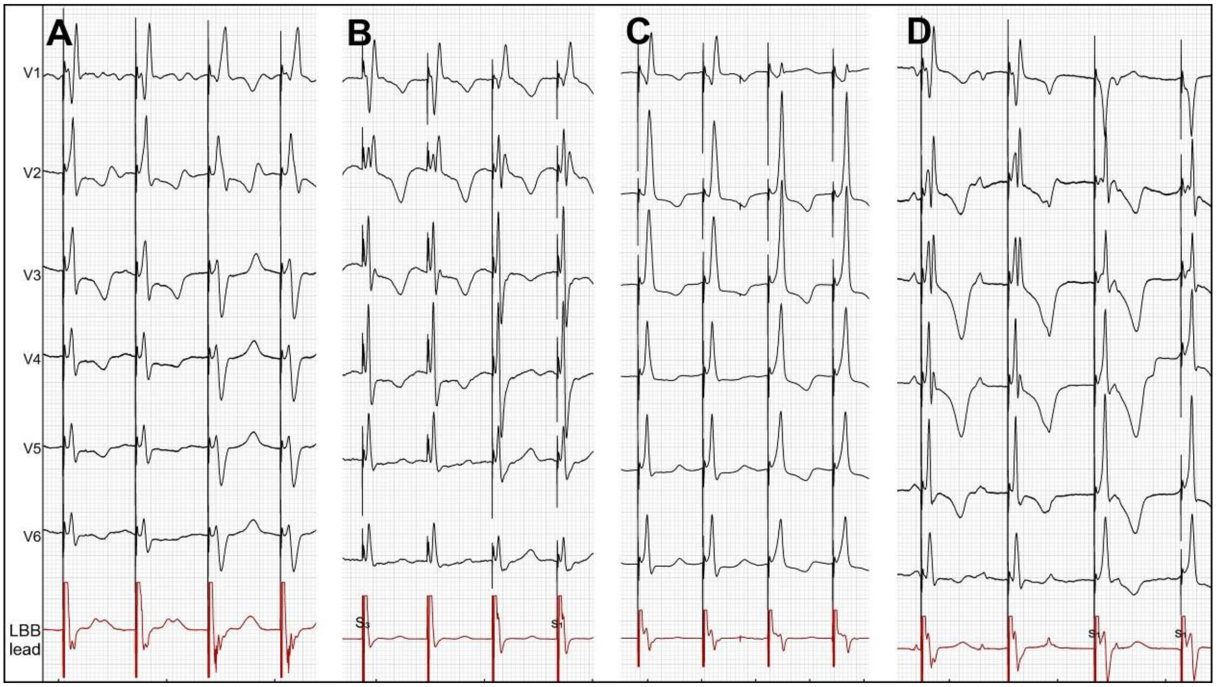


**Supplemental Figure 1.** Panels A–B: transition from non-selective to selective left bundle branch pacing (LBBP). Note the typical changes: appearance of a broad R wave in lead V1, increase in V1 R wave peak time, appearance of a deep S wave in lead V6, and emergence of a discrete, rapid component in the first 50 ms of the endocardial tracing (LBB lead). Panels C–D: transition from non-selective LBBP to left ventricular septal myocardial pacing. Note the typical changes: disappearance of or reduction in R wave amplitude in lead V1, disappearance of S wave in lead V6, QRS prolongation with prolongation of V6 R wave peak time, and delay of the maximal deflection in the endocardial signal. Sweep speed = 25 mm/s.

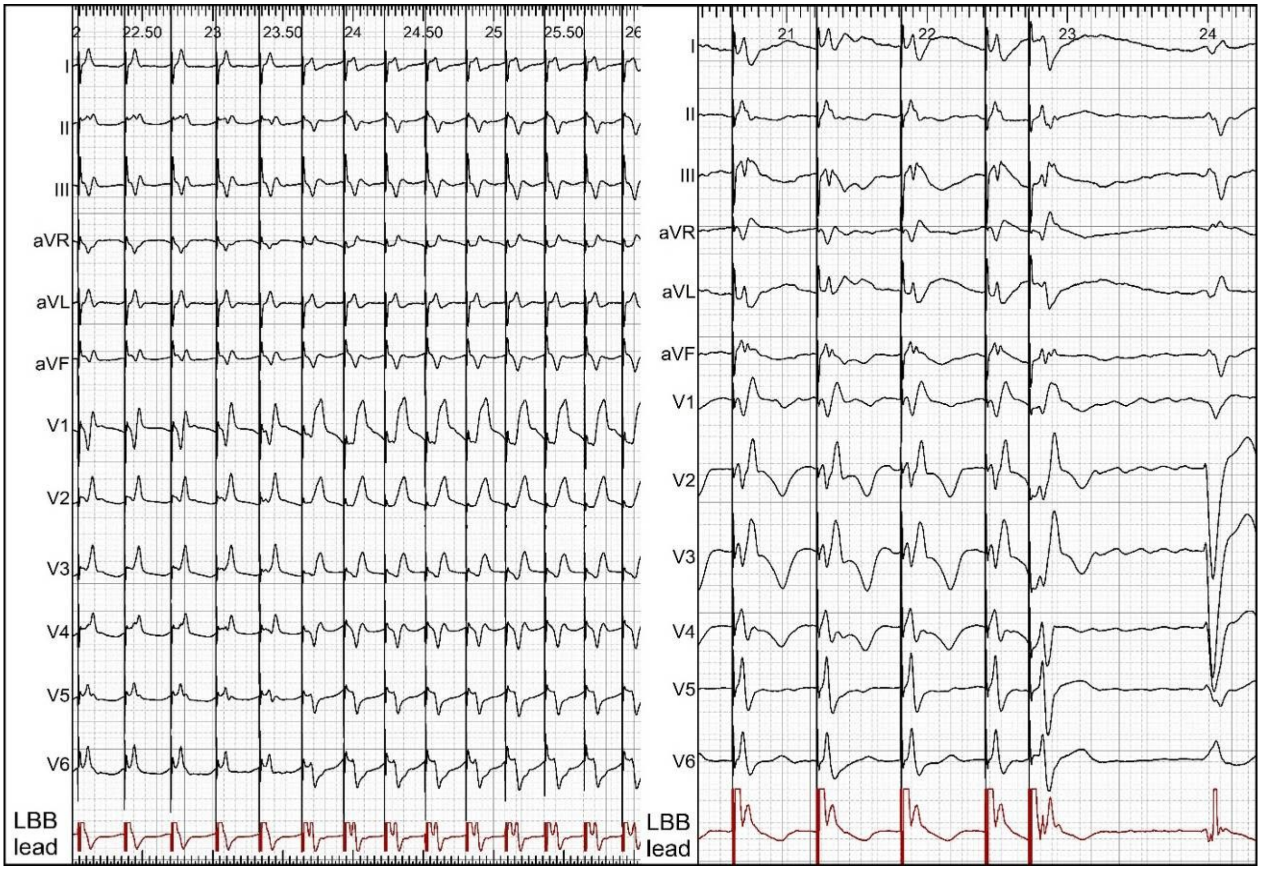


**Supplemental Figure 2**. Left panel: during incremental pacing at a cycle length of 300 ms, there was a loss of myocardial capture, resulting in transition from non-selective to selective left bundle branch pacing. Note the typical changes: appearance of a broad R wave in lead V1, increase in V1 R wave peak time, appearance of a deep S wave in leads V6 and I, and emergence of a discrete potential in the endocardial tracing (LBB lead). Right panel: during programmed pacing, the premature extrastimulus found the septal myocardium refractory, resulting in transition from non-selective to selective left bundle branch capture with similar QRS morphological changes as during the incremental pacing on the left panel; note the appearance of a discrete potential in the endocardial tracing and a clear latency interval in lead V6. Sweep speed = 25 mm/s.
